# Covid-19 Prediction in USA using modified SIR derived model

**DOI:** 10.1101/2020.12.20.20248600

**Authors:** Jathin desan

## Abstract

The Covid-19 pandemic is rapidly extended into the extraordinary crisis. Based on the SIR model and published datasets the Covid-19 spread is assessed and predicted in USA in terms of susceptible, recovered and infected in the communities is focused on this study. For modelling the USA pandemic prediction several variants have been utilized. The SIR model splits the whole population into three components such as Susceptible (S), Infected (I) and Recovered or Removed (R). A collection of differential equations have been utilized to propagate the model and resolve the disease dynamics. In the proposed study, the prediction of covid-19 based on time is performed using the modified SIR derived model SIR-D with discrete markov chain. This proposed technique analyse and forecasting the covid-19 spread in 19 states of USA. The performance analysis of the proposed Analytical results revealed that though the probable uncertainty of the proposed model provides prediction, it becomes difficult to determine the death cases in future.

## I. INTRODUCTION

COVID-19 expanded as Corona Virus Disease is an exact respiratory disease in human-beings that is caused by a virus called corona. At present there is no medicine that is clinically proven to cure this disease. Researchers have recommended that a tested and medically proven vaccine is away at a rate of one to two years. Many countries have adopted national lockdown as per the guidelines of WHO (World Health Organization) so as to reduce its spread. Formerly as per the study [1, 2] the COVID-19 case commenced in USA on 21^st^ of January, 2020 in the state of Washington. Later this particular disease spread rapidly to almost many countries once the virus has been tested in many centres. Epidemiology is a field of medicine that handles distribution, incidence and potential disease control in a population. SIR model, one of the epidemiological models [3] is employed to model the disease outspread in a population. This [4] particular model calculates the theoretical count of people affected with an infectious disease over a time in a closed population. This SIR model [5] functions by making an assumption such that there is an uninterrupted contact amongst the susceptible and infected population. This assumption is a contravention with regard to a situation where various constraints such as social distancing and quarantine have been imposed. In this, the population that have been infected is quarantined and will not contribute to the disease spread. Thus to enhance the prediction of disease outspread, this research has made several modifications in SIR model based on the existing SIR model to make predictions more accurately. On the other hand, the infected individuals have been categorized on the basis of testing which is shown in figure.1. The infected person is quarantined or not quarantined based on the results attained from testing. If the result of the particular person is positive then he/she is quarantined. Likewise if a negative result is obtained then the particular person is un-quarantined. Hence the overall predictions for the spread of the disease is possible through the use of the proposed SIR model.

**Figure.1.**
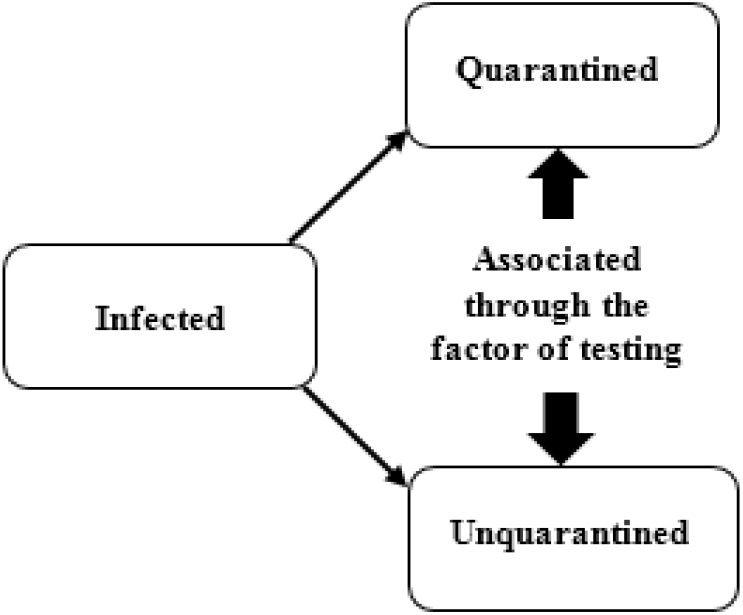
Overview of predictions for COVID-19 infected person [4]

Based on the covid-19 prediction and SIR model, the major contribution of this study involves,

- To predict the COVID-19 probability distribution in USA in terms of time/days using the discrete Markov chain.
- To evaluate the infected population by utilizing modified SIR derived model (Susceptible, Infected, and Recovered) SIR-D.
- To analyse the predictable results from the performance analysis of the proposed SIR-D model.

### 1.1 Paper Organization

The following section 2 describes the literature review COVID-19 probability distribution in USA. Further section 3 illustrates the proposed methodology, followed by results and discussion briefly explained in the section 4. Finally the paper is concluded in section 5.

## II. RELATED WORK

The following section deals with the different schemes, for the covid-19 pandemic prediction in various countries, The COVID-19 (Corona Virus Disease) was initially stated at 2019 by the end of the year and it has been spreading rapidly to nearly all countries. It has extreme infectious nature. Due to this impact, most of the countries have made compulsory measures to manage its spread. This article has used the infectious data in the early phase to discuss certain parameters such as average duration of the infectious period, determination of the fundamental reproduction number and assessment of the peak timing of the epidemic wave. Day-to-day fatalities and case reports for many countries over a certain period has been assessed through the use of SIR (Susceptible-Infected-Removed) model. The proposed model has been evaluated by conducting a comparative analysis in which the actual data and the predictions has been compared. The analytical results revealed that the predictions have been appreciated for all countries excluding USA as far as measures for lock-down were maintained [6]. Additionally, [7] Conducted a study on a Euclidean network regarding the day-to-day COVID-19 cases as well as cumulative data in China. The cumulative data is attained from a SIR model and can be made suitable to an empirical form. It has been stated that this SIR model can replicate with efficient accuracy for certain metric values. It can also detect the time at which the particular epidemic will be over. Similarly [8] assessed the SIR model for COVID-19. The transmission rate is varied across many countries and excels the recovery rate that permits a fast spread. Various analysis of this study has been conducted. From the analysis it has been concluded that the spread of COVID-19 occurs except three countries that is japan, china and Korea. Further [9] examined the pandemic of COVID-19 with the use of machine learning techniques and SIR model for forecasting. Beneath the optimistic estimation regarding the pandemic of corona in few countries will come to an end soon. It has also been found that for many part of the countries, the anti-pandemic hit will occur by the April end. Here [10] Investigated the pandemic of COVID-19 particularly in Sweden. SIR and SI (Susceptible-infected) model have been utilized for analysis. The proposed model replicate the infected cases and provides similar predictions. Analytical results revealed that though the probable uncertainty of the proposed model provides prediction, it becomes difficult to determine the death cases in future. Thus policy response has been developed [11] by introducing three models on the basis of data driven for COVID-19 by means of minimum parameters to give visions into the disease spread that might be utilized for policy responses development. The three models that have been involved in the study include a self-exciting branching which is a process model, exponential growth and SIR model. The proposed three models have been associated quantitatively. The SIR model have been utilized for potential-effects demonstration with respect to distancing measures in short-term in the US (United States).

Moreover [12] Studied the detection issues in a network where the information spread tracks the famous SIR model. Here an assumption has been made in the undertaken simulation where all the nodes are initially in the susceptible state in the network except the information source. The information source has been assumed to be in the infected state. Thus the simulation results revealed that, the estimator provided by the algorithm called reverse-infection (RI) is nearer to the actual source for tree networks. Further evaluation has been executed to assess the RI performance on various real world networks. Likewise [13] attained an accurate analytical solution of SIR model in a parametric form. The major characteristics of the accurate solution have been numerically examined and it has been revealed that it replicates accurately the numerical result of the model equations. The accurate solution has been found to be significant as biologists could utilize it to run experiments to detect the outspread of COVID-19 by presenting natural conditions. Hence this study can be utilized to learn the several ways to manage the epidemic spread. Further [14] introduced an approach on the basis of α – path that can regulate the proposed SIR model. This approach has been utilized to compute the uncertainty distributions and pertained Expected Values (EV) of the solutions. The results specify that the lockdown policy accomplishes hundred percent efficiency after thirteenth of February in the year 2020 that is constant with the existing studies.

The proposed system creates new opportunities to solve multi-dimensional doubtful new applications and differential equations in other fields. Hence this paper [15] used Gaussian mixture model for prediction of COVID-19. The proposed system has been analysed to determine the performance and efficiency in terms of prediction. The analytical results have shown that the proposed system provides optimistic results with the ability to function as a better supplement to the existing methods for monitoring the predictions of COVID-19 pandemic continuously. Further the spread of COVID-19 within a community has been examined through the [16] Proposed SIR model that affords a theoretical framework. Analysis has been performed by considering the period from January to June of 2020. This particular period consisted data previously and at the time of strict measure implementation. Various predictions have been proposed based on certain parameters. Comparative analysis of the recorded data with the actual data has been performed to determine the spread of this particular disease. The results from comparative analysis revealed that the disease spread can be managed in all communities if strong policies and correct restrictions are employed to manage the infection rates before the disease outspread. Similarly an another model called Poisson model has been developed [17] by means of removal rates and transmission with respect to time-dependency to account for potential random errors while computing and reporting a reproduction number with respect to time-dependent disease. This might reflect the efficiency of the virus management approaches. The proposed method can be employed to review the pandemic in many severely affected countries. The forecast and analysis has been performed regarding the growing outspread of the disease. An interactive web application has also been developed to help the readers to use the proposed method. Various other analytical tools has also been presented [18] for prediction, planning and forecasting at the time of pandemic. Prognosticated the growth rate of COVID-19 with epidemiological, statistical, a new forecasting technique and deep and machine learning models on the basis of clustering and nearest neighbours. Additionally forecasting has been performed with regard to abundant demand for services and products at the time of pandemic through the use of simulations (decisions of government regarding lockdown) and auxiliary data. It has been found that the results can aid planners and policy makers to take better decisions in the course of future and ongoing pandemics. Thus many forecasting models is yet to be identified.

Although [19] many theoretical methods have been proposed for predicting the threshold of epidemics that includes QMF (Quenched-Mean Field), MFL (Mean-Field Like) and DMP (Dynamical Message Passing) method. Once these methods have been employed for epidemic threshold prediction they frequently provide different outcomes and its accuracy level has also been found to unknown. These challenges has been consistently analyzed between accuracy level and different outcomes by examining and studying the SIR model on configuration networks that is uncorrelated. It has been found that the performance of MFL method has been better than QMF and DMP. Several transactional datasets has also been incorporated in [20] to examine the role of policies of social distancing in many countries. The study has also reviewed the pandemic of coronavirus disease and its transmission rate over a period of five weeks. This process depend on the official reports of COVID-19 and SIR model. Simulation has been carried to assess the system performance. The simulation results revealed the concerning of social distancing envelopments has been found to be effective in reducing the COVID-19 spread. Few models have also been designed to predict the pandemic of COVID-19. [21] This study has incorporated the (EPM) Epidemic Prediction Model with respect to real pandemic data globally and has taken into account the execution of control measures and the impact of environmental factors to create a prediction system globally. Thus the proposed study predicts the disease (COVID-19) over globe day by day in each country. Similarly in Brazil [22] Demonstrated and forecast the early commencement of the pandemic of COVID-19 by utilizing the recent Brazilian data. This study has used SIR model with two variations and comprised the influence of measures in social distancing. Hence the results exhibited that the policy of social distancing executed by the government is capable of reducing the infection pattern of COVID-19. Likewise [23] COVID-19 has caused pandemic situations and an elaborate analysis of this study is yet to come. But at the same time an evaluation regarding the disease parameters has to be executed to estimate early predictions about the possible durations for the sustaining pandemic. For this purpose SIR model has been proposed to make early estimations about the pandemic. It has been found that the prediction accuracy diminishes when the isolation speed gets altered. Thus measures for lockdown of this particular disease can be implemented to reduce the spread. For this purpose [24] Employed SIR model to detect and predict the various pandemic of COVID-19. This study has explored the rate of infection spread and aids doctors to take decisions accordingly to prevent the spread of disease.

## III. Proposed Methodology

The proposed system comprised with two sections for predicting the COVID-19 probability distribution in time using the discrete Markov chain in USA and another section, the infected population is evaluation in time and by utilizing SIR differential equation model (Susceptible, Infected, Recovered) the information is retrieved. Based on the standard SIR model the proposed study derived a mathematical model. For evaluating the infectious disease outbreaks, SIR model is considered as fundamental statistical tool. However, it is not considering the asymptomatic cases which are the infection source in the whole population and also infection transmitted through the touching objects. With respect to disease transmission, various random factors are presented. By considering all these factors, the proposed study developed the modified SIR derived (SIR-D) model. For SIR-D, the parameter estimation is applied to the time-series data related with covid-19 infection rate in USA which define its impacts.

### 3.1 Discrete Markov chain

A set of random variables as X0, X1, X2… sequence is described as discrete-time Markov chain with the Markov property specifically that the probability of moving to following state based on the current state and not of on past states. In this study, the discrete markov chain comprised of 19 states which representing the USA political territory division, hence, *D*={*d*:*d* ∈ *U,i*, ∈ ℕ} United States is U, it is mapped into discrete states set *S*_*t*=_{*s*_*t*1,_ *s*_*t*2,_ ….*s*_*t*32_} of *f:D→ S*_*t*_ *in* bijective style. The bijective function is the one-to-one representation or otherwise refer as invertible function defined as function among the two sets elements in which every element in which every element in one set is paired with precisely one element of initial set.

Let consider V as the process, followed as *T*_*e*_, ⊂ V, 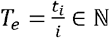, [i] = [day] or *i*–i th day, the temporal parameter equivalent unit is day. For each relationship existence among a-th and k-th state, the probability conditions are developed as ∀*s*_*ta*_, *R*_*stk*_,∃.𝒫_*ak*_. For the transition from ath to kth state, the associated probability is exists and it is maintained among all possible interactions with a-th and k-th state. For every *s*_*ta*_*R*_*stk*_, the associative probability is 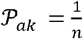, a-th state is interrelated with n various states. By using the territorial boundaries sharing, the interaction among the one state and another one is exhibited. By using the stochastic matrix*P*_*m*_ 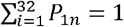,

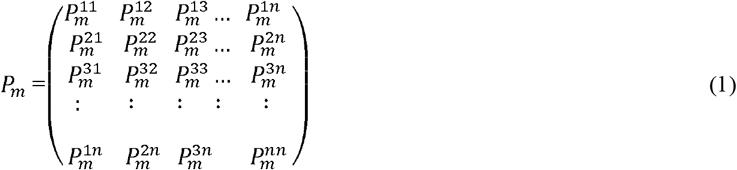

By using the corresponding report from Jan 22. 2020 to April 14, 2020 by USA health department, the first vector exhibited the features of covid-19 with cases from 19 states in USA. The cardinality of the set of cases B as the total amount of cases, |B| = 11 and for the given state 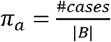 the initial vector probabilities π ^0^. For discrete time *t*_*a*_ ∈ *T*_*e*_,

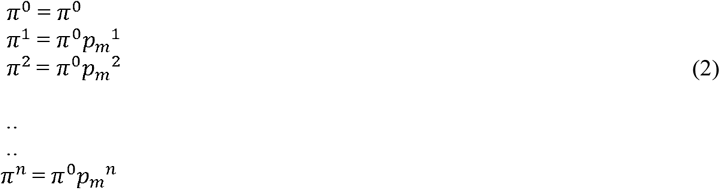

The stochastic powers have been computed proceeded to the probability distribution when n = 2, 4, 8 ….320 defines that [i] = [day] as temporal parameters and the covid-19 probability distributions for following dates are, from Jan 22. 2020 to April 14, 2020 by USA health department.

### 3.2 Modified SIR derived (SIR-D) model

From the set of population, the susceptible, infected and recovered amount are identified from the proposed SIR derived (SIR-D) model. Form the below expression, this proposed model is derived,

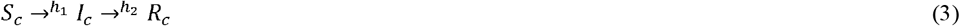

From Eq. (3), the population of susceptible is *S*_*c*_, infected is *I*_*c*_ and recovered is *R*_*c*_ through chemical kinetics and to these sets, the analogy is generated as,

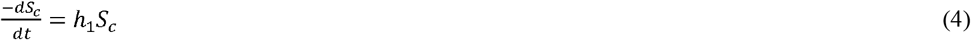

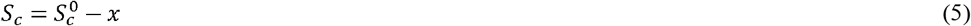

From Eq. (5), initial susceptible population is 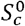, lost of susceptible people amount at the disease’s time is S_c_™ x

Otherwise,

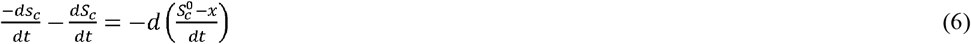

The represented differential equation,

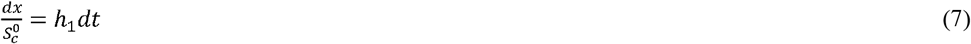

The final expression integrated as,

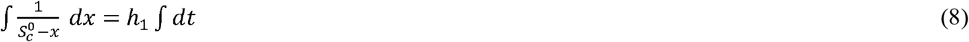

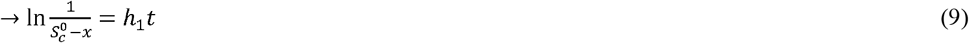

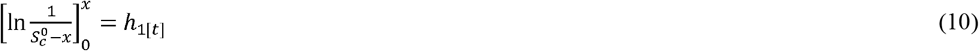

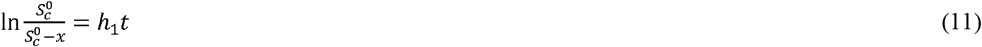

The following information is obtained further,

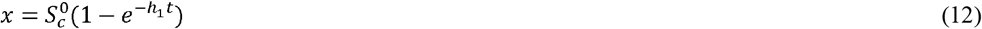

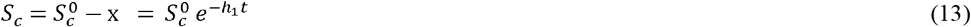

The infected people is computed and equalities are defined as,

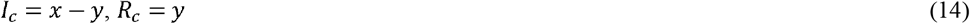

Further,

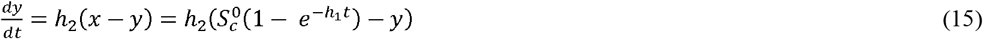

It is the linear differential equation of non-homogenous as,

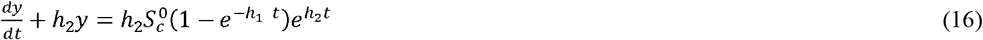

The integrating factor introduced,

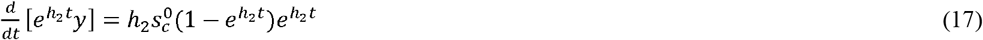

Integrated as,

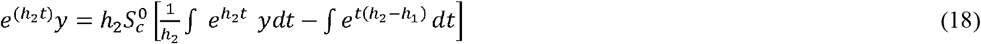

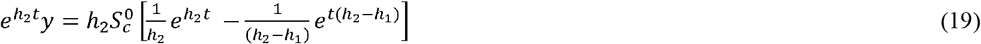

The limits are substituted as,

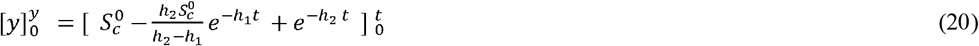

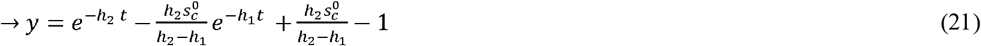

Recalling,

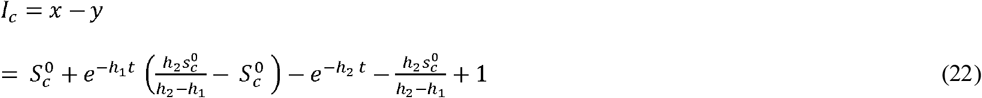

The initial susceptible population is,

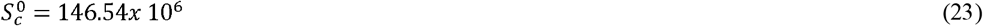

The constants are,

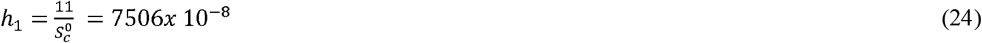

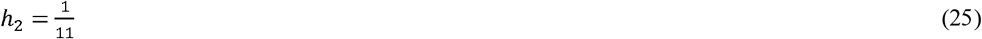

## IV. Results and Discussion

In this proposed study, the covid-19 prediction is performed by using the modified SIR derived model SIR-D with discrete markov chain. The performance analysis results are depicted in the following section

### 4.1 Dataset Description

For covid-19 prediction and analysis, the dataset is collected from Covid-19 confirmed cases from US. The number of confirmed cases, number of recovered cases and number of susceptible cases are collected from Jan 22, 2020 to April 14, 2020 considered as proposed dataset. The dataset is collected from 19 states in USA. The source of the dataset link is https://data.world/shad/covid-19-time-series-data.

### 4.2 Performance analysis

The cases of COVID-19 in Alabama is shown in figure 2. Here the susceptible case is found to be in peak initially, but as time and days passed the rate of susceptible cases gradually decreased. The infected rate is found to be moderate and this rate also gradually decreased as days and time passed. The recovered rate of COVID-19 cases is found to be zero initially, that is the susceptible and recovered rate has been found to be inversely proportional. It has been predicted that the recovery rate gradually increased and reached the peak when the days passed.

**Figure.2.**
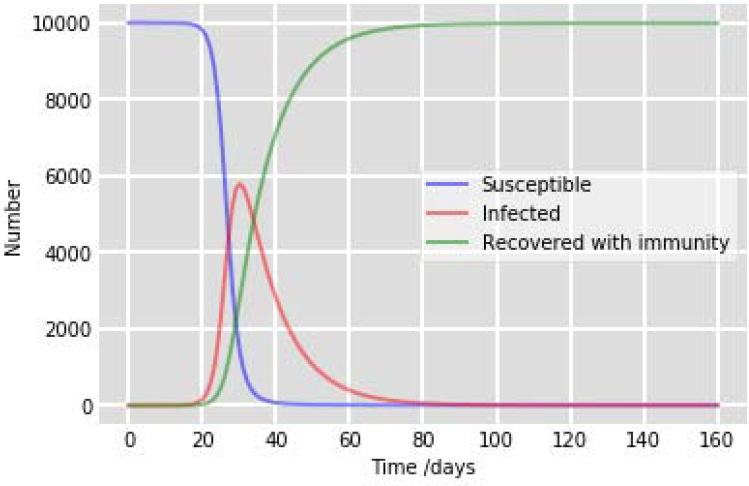
Different cases of COVID-19 in Alabama.

The cases of COVID-19 in California is shown in figure 3. Initially the susceptible case is found to be in peak, but as time and days passed the rate of susceptible cases decreased gradually. The infected rate is found to be moderate and this rate also gradually decreased as days and time passed. The susceptible and recovered rate has been found to be inversely proportional, that is the recovered rate has been found to be zero when the susceptible rate has been found to be high and vice-versa. Thus it has been accurately predicted that the recovery rate gradually increased and reached the peak as the days passed.

**Figure.3.**
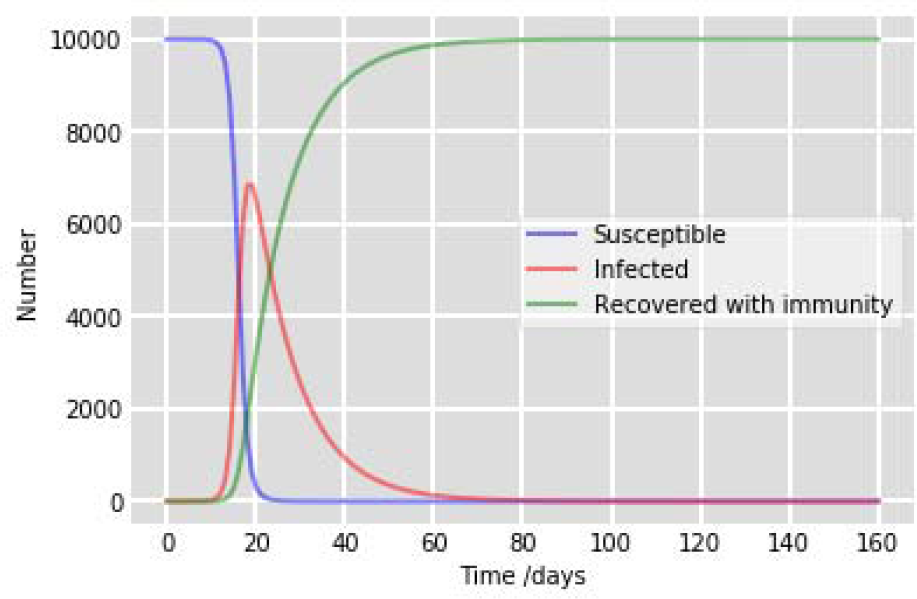
Different cases of COVID-19 in California.

The cases of COVID-19 in Colorado is shown in figure 4. Here the susceptible case is found to be in peak initially, but as time and days passed the rate of susceptible cases gradually decreased. The infected rate is found to be moderate and this rate also gradually decreased as days and time passed. The recovered rate of COVID-19 cases is found to be zero initially, that is the susceptible and recovered rate has been found to be inversely proportional. It is predicted that the recovery rate gradually increased and reached the peak when the days passed.

**Figure.4.**
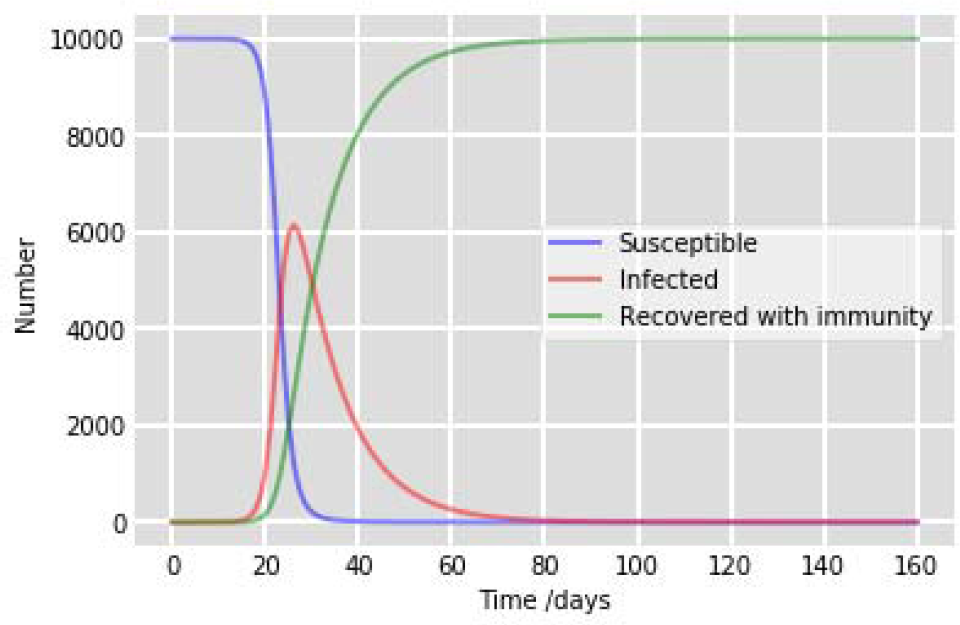
Different cases of COVID-19 in Colorado.

The cases of COVID-19 in Florida is shown in figure 5. Initially the susceptible case is found to be in peak, but as time and days passed the rate of susceptible cases decreased gradually. The infected rate is found to be moderate and this rate also gradually decreased as days and time passed. The susceptible and recovered rate has been found to be inversely proportional, that is the recovered rate has been found to be zero when the susceptible rate has been found to be high and vice-versa. Thus it has been accurately predicted that the recovery rate gradually increased and reached the peak as the days passed.

**Figure.5.**
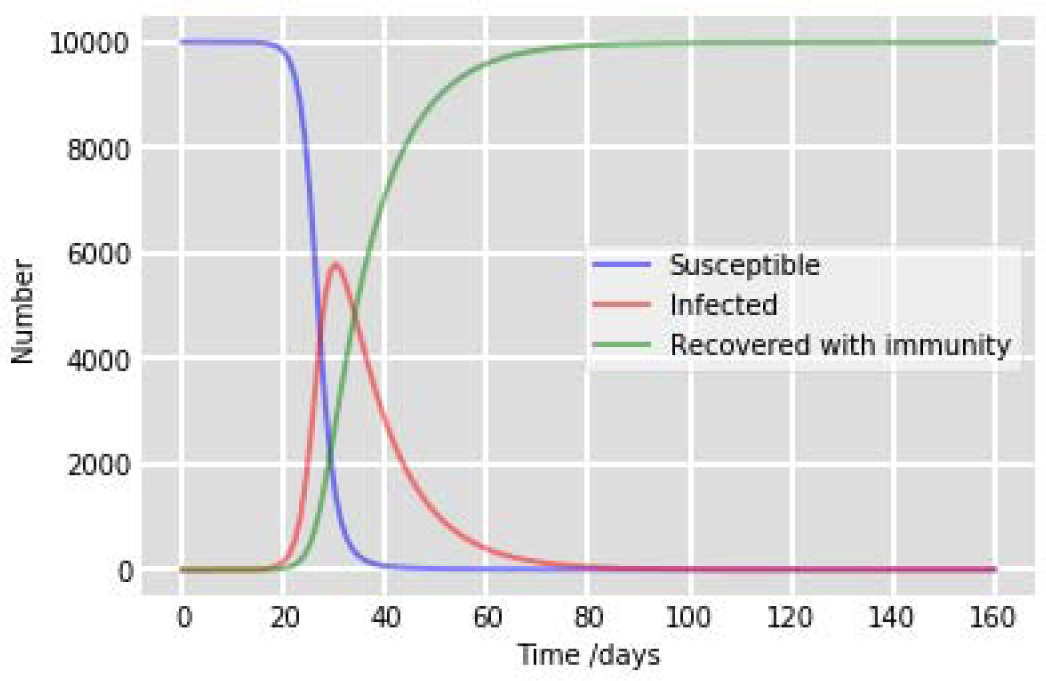
Different cases of COVID-19 in Florida.

The cases of COVID-19 in Hawaii is shown in figure 6.Here the susceptible case is found to be in peak initially, but as time and days passed the rate of susceptible cases gradually decreased. The infected rate is found to be moderate and this rate also gradually decreased as days and time passed. The recovered rate of COVID-19 cases is found to be zero initially, that is the susceptible and recovered rate has been found to be inversely proportional. It is predicted that the recovery rate gradually increased and reached the peak when the days passed.

**Figure.6.**
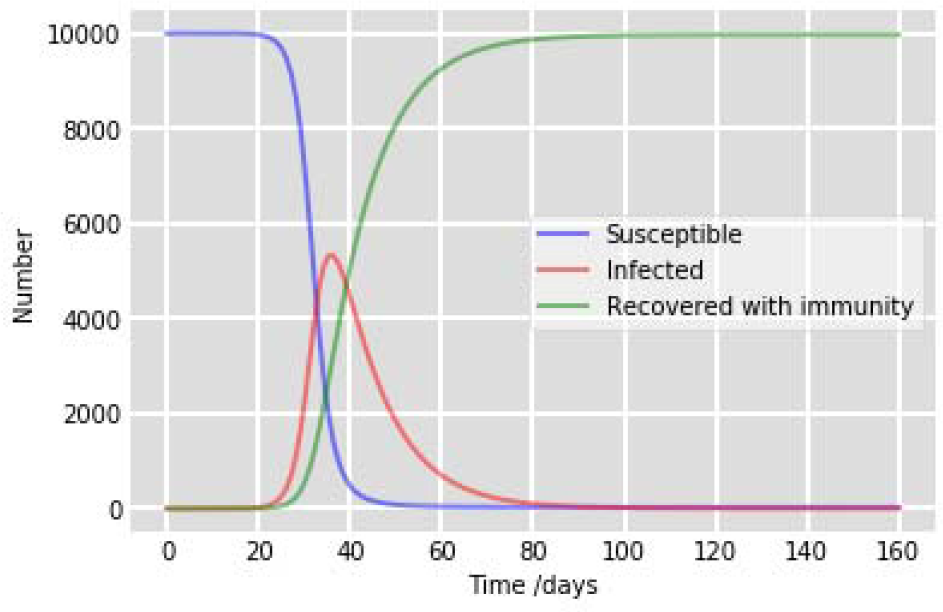
Different cases of COVID-19 in Hawaii.

The cases of COVID-19 in Illinois is shown in figure 7. Initially the susceptible case is found to be in peak, but as time and days passed the rate of susceptible cases decreased gradually. The infected rate is found to be moderate and this rate also gradually decreased as days and time passed. The susceptible and recovered rate has been found to be inversely proportional, that is the recovered rate has been found to be zero when the susceptible rate has been found to be high and vice-versa. Thus it has been accurately predicted that the recovery rate gradually increased and reached the peak as the days passed.

**Figure.7.**
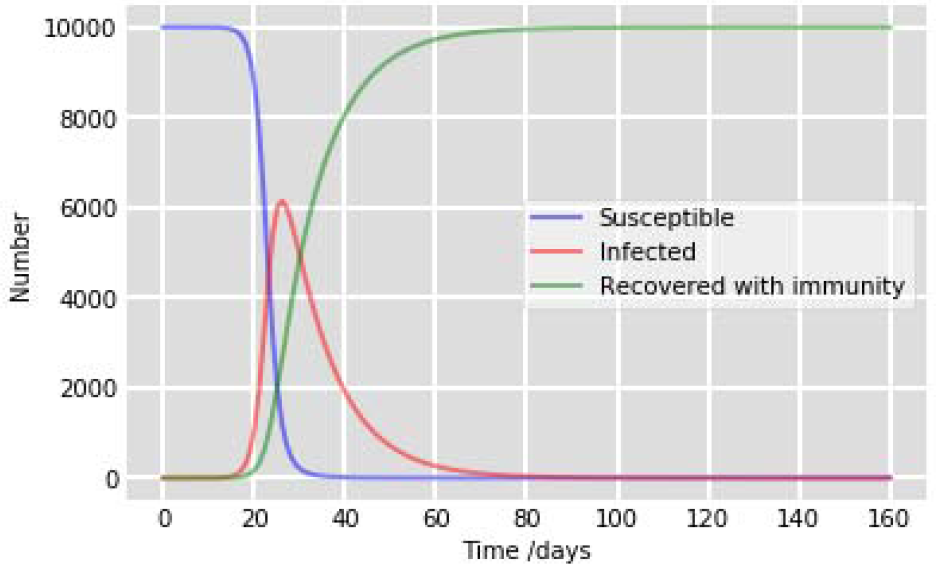
Different cases of COVID-19 in Illinois.

The cases of COVID-19 in Massachusetts is shown in figure 8. Here the susceptible case is found to be in peak initially, but as time and days passed the rate of susceptible cases gradually decreased. The infected rate is found to be moderate and this rate also gradually decreased as days and time passed. The recovered rate of COVID-19 cases is found to be zero initially, that is the susceptible and recovered rate has been found to be inversely proportional. It is predicted that the recovery rate gradually increased and reached the peak when the days passed.

**Figure.8.**
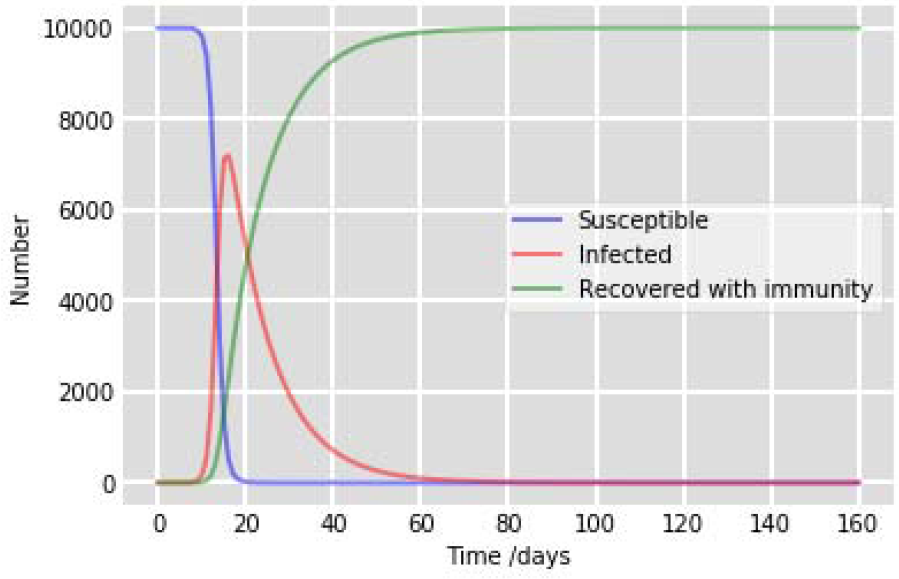
Different cases of COVID-19 in Massachusetts.

The cases of COVID-19 in Michigan is shown in figure 9. Initially the susceptible case is found to be in peak, but as time and days passed the rate of susceptible cases decreased gradually. The infected rate is found to be moderate and this rate also gradually decreased as days and time passed. The susceptible and recovered rate has been found to be inversely proportional, that is the recovered rate has been found to be zero when the susceptible rate has been found to be high and vice-versa. Thus it has been accurately predicted that the recovery rate gradually increased and reached the peak as the days passed.

**Figure.9.**
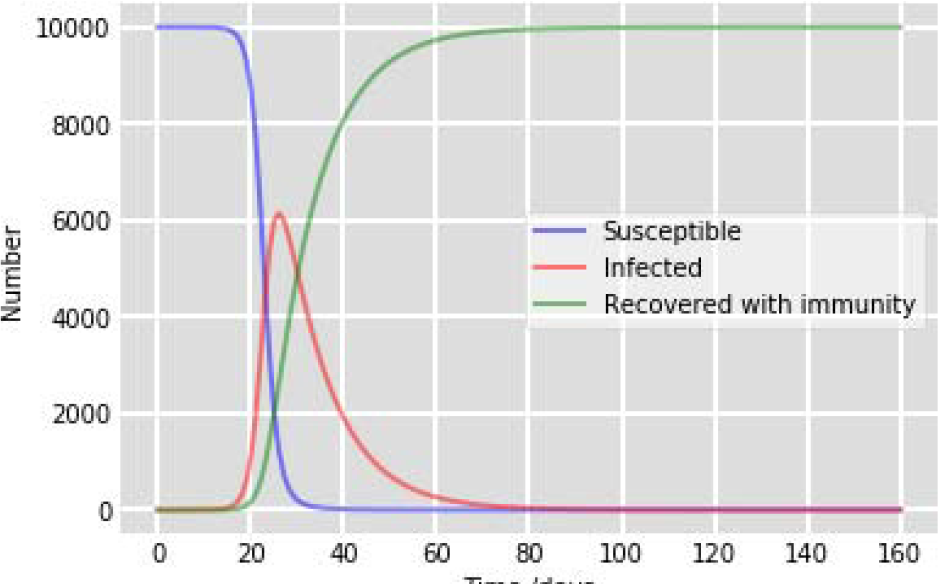
Different cases of COVID-19 in Michigan.

The cases of COVID-19 in Missouri is shown in figure 10. Initially the susceptible case is found to be in peak, but as time and days passed the rate of susceptible cases decreased gradually. The infected rate is found to be moderate and this rate also gradually decreased as days and time passed. The susceptible and recovered rate has been found to be inversely proportional, that is the recovered rate has been found to be zero when the susceptible rate has been found to be high and vice-versa. Thus it has been accurately predicted that the recovery rate gradually increased and reached the peak as the days passed.

**Figure.10.**
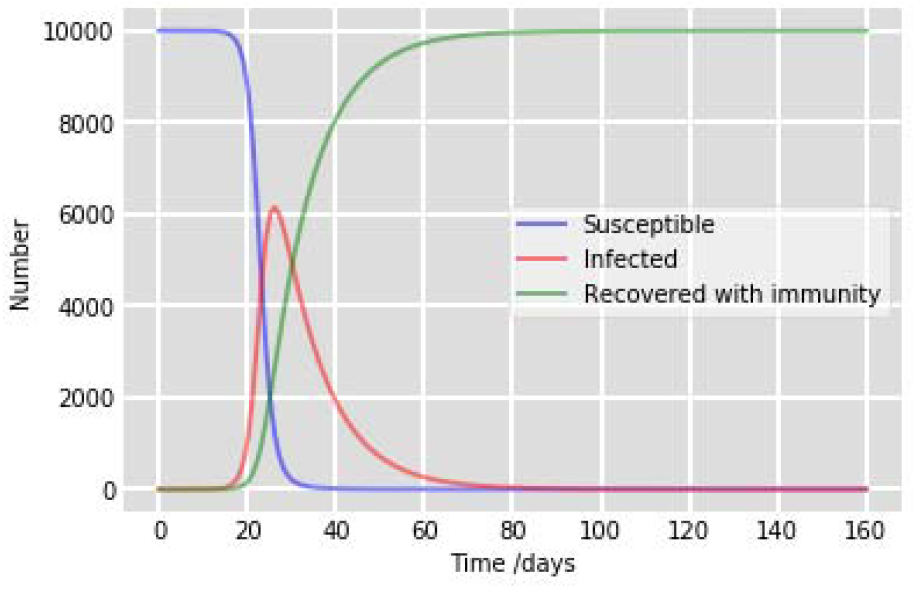
Different cases of COVID-19 in Missouri.

The cases of COVID-19 in New Jersey is shown in figure 11.Here the susceptible case is found to be in peak initially, but as time and days passed the rate of susceptible cases gradually decreased. The infected rate is found to be moderate and this rate also gradually decreased as days and time passed. The recovered rate of COVID-19 cases is found to be zero initially, that is the susceptible and recovered rate has been found to be inversely proportional. It is predicted that the recovery rate gradually increased and reached the peak when the days passed.

**Figure.11.**
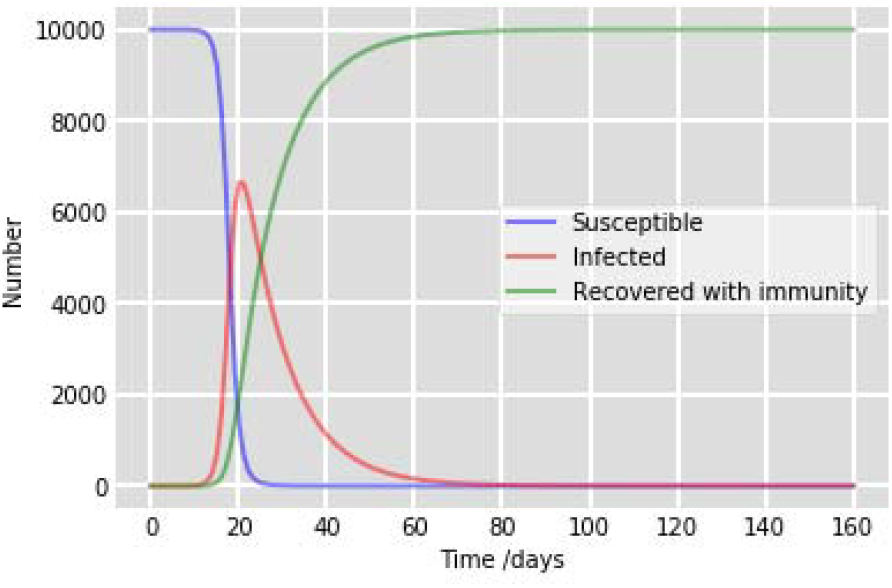
Different cases of COVID-19 in New Jersey.

The cases of COVID-19 in New Mexico is shown in figure 12. Initially the susceptible case is found to be in peak, but as time and days passed the rate of susceptible cases decreased gradually. The infected rate is found to be moderate and this rate also gradually decreased as days and time passed. The susceptible and recovered rate has been found to be inversely proportional, that is the recovered rate has been found to be zero when the susceptible rate has been found to be high and vice-versa. Thus it has been accurately predicted that the recovery rate gradually increased and reached the peak as the days passed.

**Figure.12.**
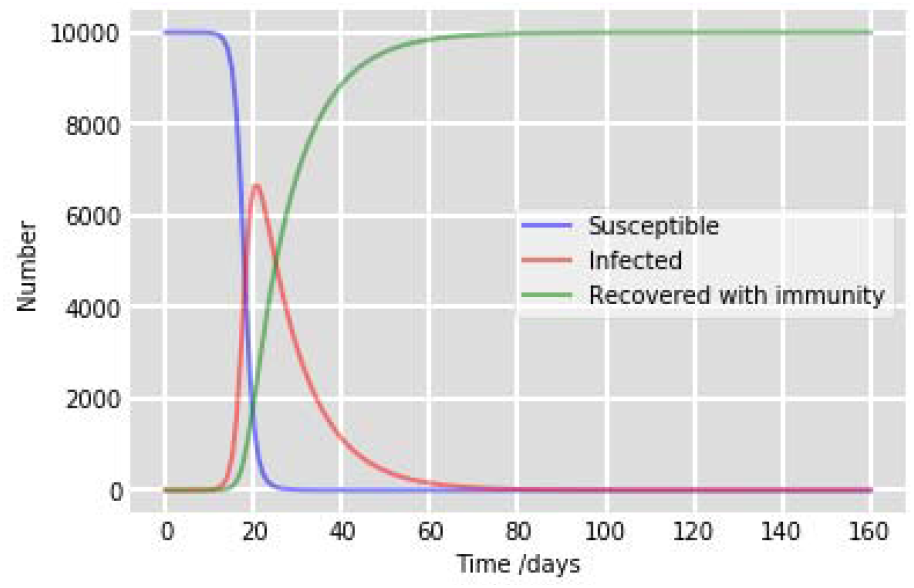
Different cases of COVID-19 in New Mexico.

The cases of COVID-19 in North Carolina is shown in figure 13. Here the susceptible case is found to be in peak initially, but as time and days passed the rate of susceptible cases gradually decreased. The infected rate is found to be moderate and this rate also gradually decreased as days and time passed. The recovered rate of COVID-19 cases is found to be zero initially, that is the susceptible and recovered rate has been found to be inversely proportional. It is predicted that the recovery rate gradually increased and reached the peak when the days passed.

**Figure.13.**
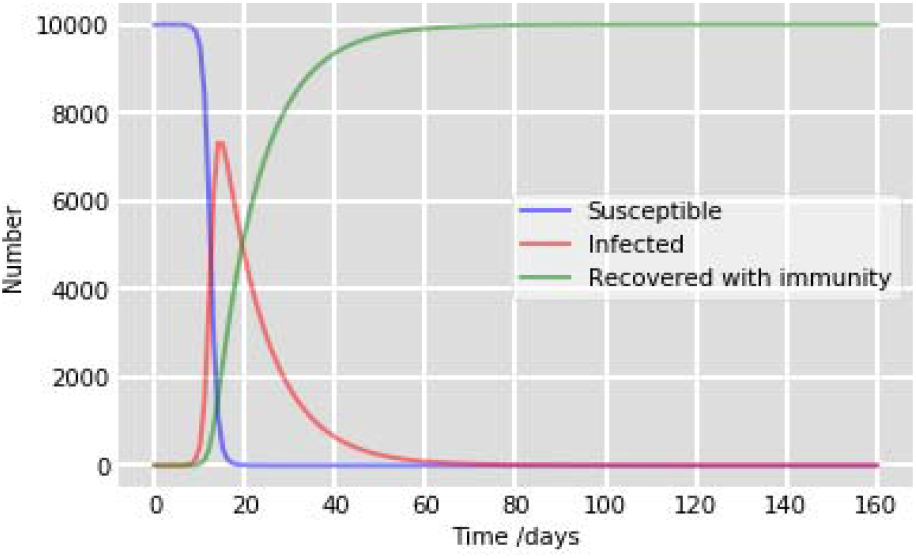
Different cases of COVID-19 in North Carolina.

The cases of COVID-19 in Ohio is shown in figure 14. Initially the susceptible case is found to be in peak, but as time and days passed the rate of susceptible cases decreased gradually. The infected rate is found to be moderate and this rate also gradually decreased as days and time passed. The susceptible and recovered rate has been found to be inversely proportional, that is the recovered rate has been found to be zero when the susceptible rate has been found to be high and vice-versa. Thus it has been accurately predicted that the recovery rate gradually increased and reached the peak as the days passed.

**Figure.14.**
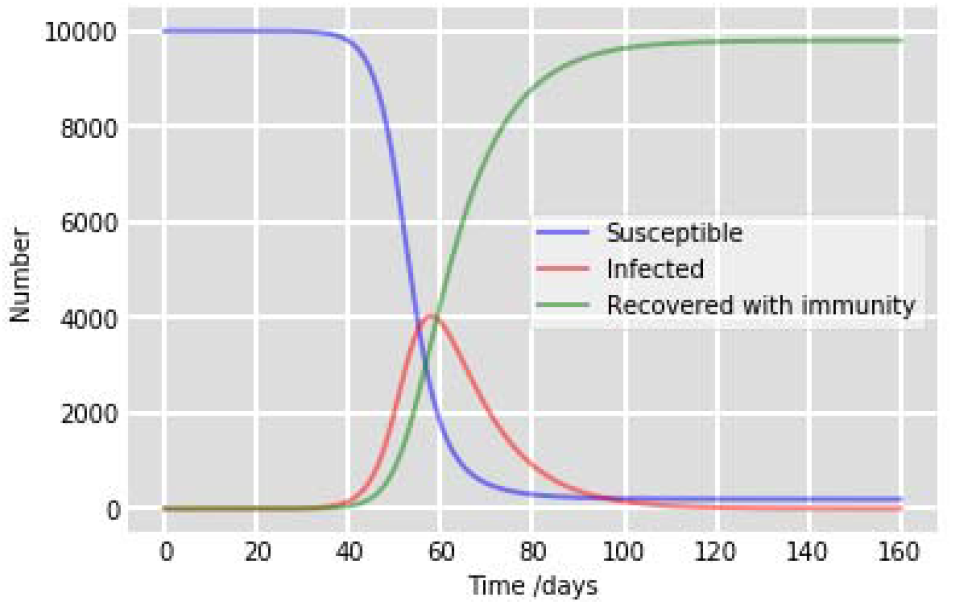
Different cases of COVID-19 in Ohio.

The cases of COVID-19 in Pennsylvania is shown in figure 15. Here the susceptible case is found to be in peak initially, but as time and days passed the rate of susceptible cases gradually decreased. The infected rate is found to be moderate and this rate also gradually decreased as days and time passed. The recovered rate of COVID-19 cases is found to be zero initially, that is the susceptible and recovered rate has been found to be inversely proportional. It is predicted that the recovery rate gradually increased and reached the peak when the days passed.

**Figure.15.**
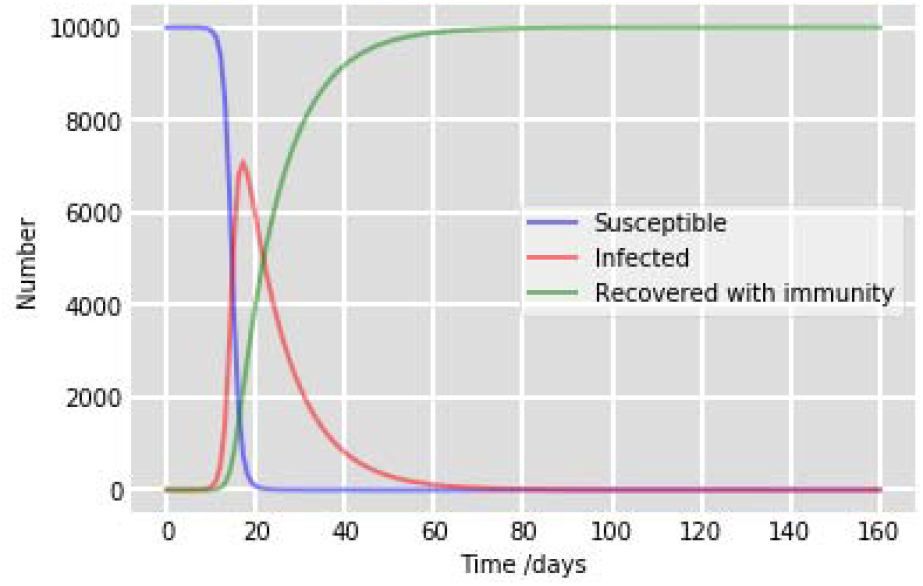
Different cases of COVID-19 in Pennsylvania.

The cases of COVID-19 in South Carolina is shown in figure 16. Initially the susceptible case is found to be in peak, but as time and days passed the rate of susceptible cases decreased gradually. The infected rate is found to be moderate and this rate also gradually decreased as days and time passed. The susceptible and recovered rate has been found to be inversely proportional, that is the recovered rate has been found to be zero when the susceptible rate has been found to be high and vice-versa. Thus it has been accurately predicted that the recovery rate gradually increased and reached the peak as the days passed.

**Figure.16.**
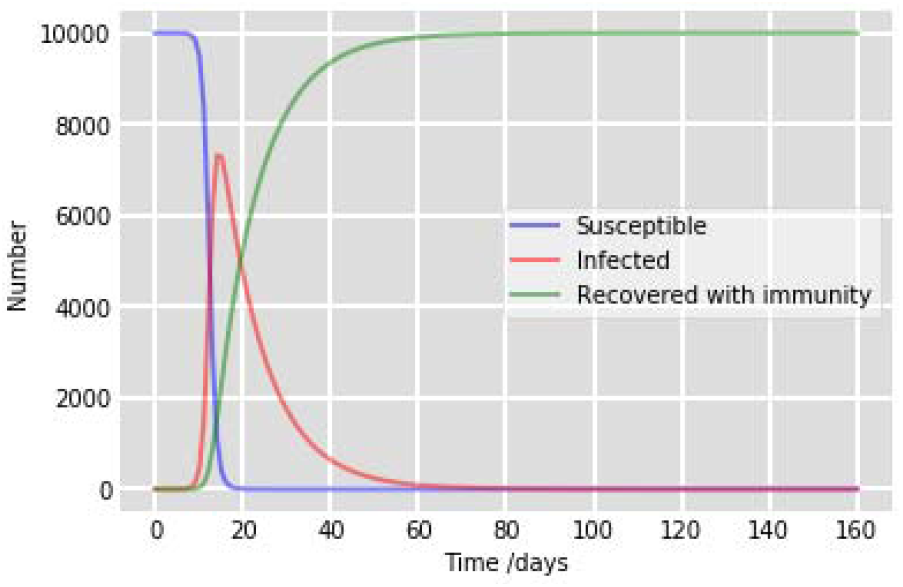
Different cases of COVID-19 in South Carolina.

The cases of COVID-19 in Texas is shown in figure 17. Here the susceptible case is found to be in peak initially, but as time and days passed the rate of susceptible cases gradually decreased. The infected rate is found to be moderate and this rate also gradually decreased as days and time passed. The recovered rate of COVID-19 cases is found to be zero initially, that is the susceptible and recovered rate has been found to be inversely proportional. It is predicted that the recovery rate gradually increased and reached the peak when the days passed.

**Figure.17.**
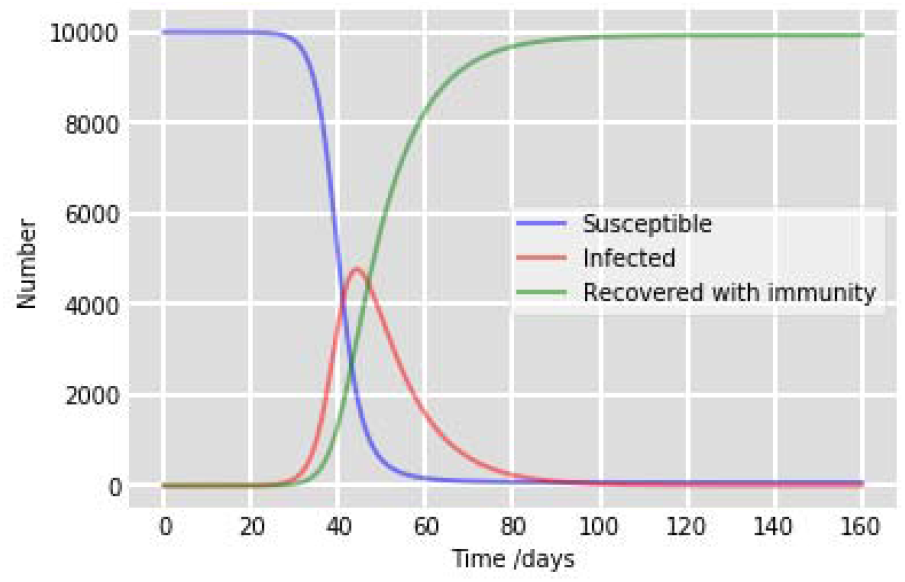
Different cases of COVID-19 in Texas.

The cases of COVID-19 in Virginia is shown in figure 18. Initially the susceptible case is found to be in peak, but as time and days passed the rate of susceptible cases decreased gradually. The infected rate is found to be moderate and this rate also gradually decreased as days and time passed. The susceptible and recovered rate has been found to be inversely proportional, that is the recovered rate has been found to be zero when the susceptible rate has been found to be high and vice-versa. Thus it has been accurately predicted that the recovery rate gradually increased and reached the peak as the days passed.

**Figure.18.**
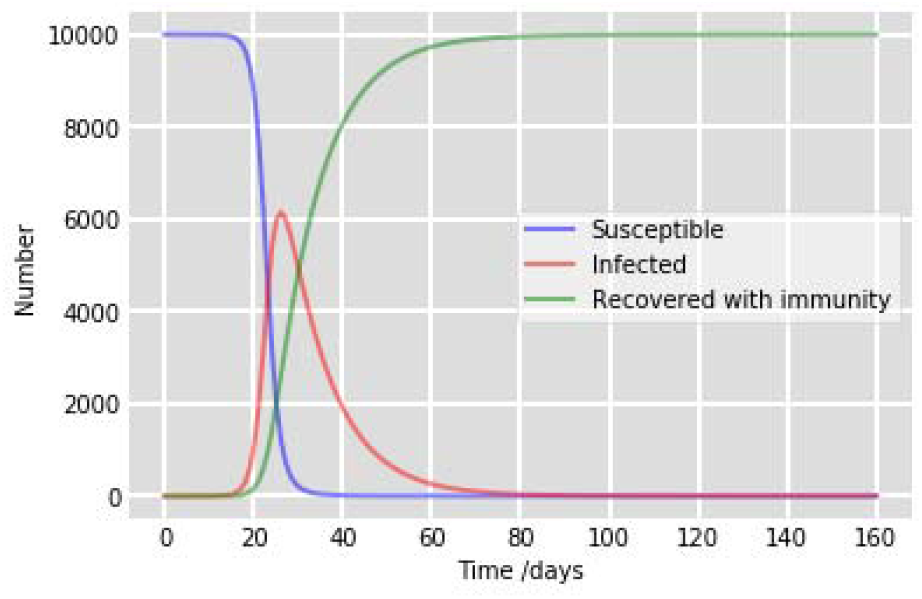
Different cases of COVID-19 in Virginia.

The cases of COVID-19 in Washington is shown in figure 19. Here the susceptible case is found to be in peak initially, but as time and days passed the rate of susceptible cases gradually decreased. The infected rate is found to be moderate and this rate also gradually decreased as days and time passed. The recovered rate of COVID-19 cases is found to be zero initially, that is the susceptible and recovered rate has been found to be inversely proportional. The recovery rate gradually increased and reached the peak when the days passed which is accurately predicted through the proposed system.

**Figure.19.**
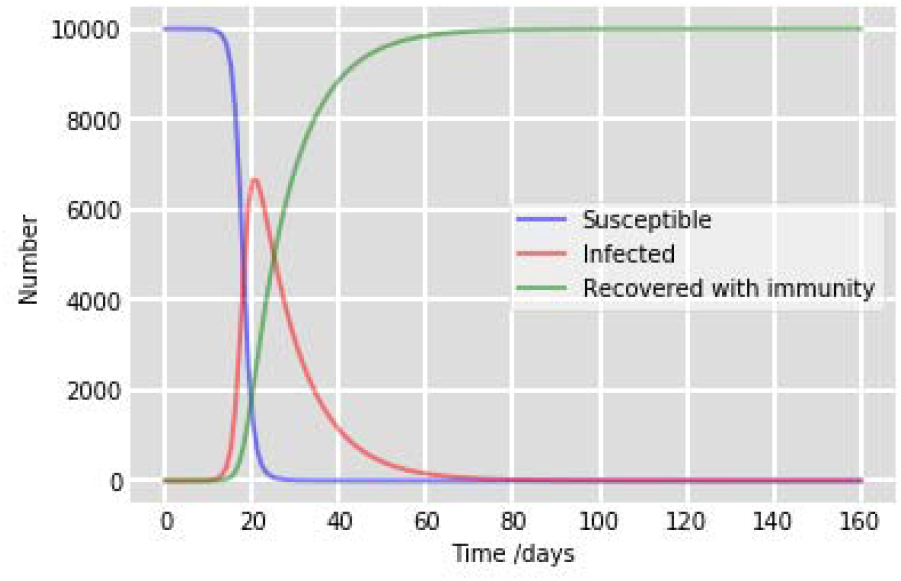
Different cases of COVID-19 in Washington.

The cases of COVID-19 in Wyoming is shown in figure 20. Initially the susceptible case is found to be in peak, but as time and days passed the rate of susceptible cases decreased gradually. The infected rate is found to be moderate and this rate also gradually decreased as days and time passed. The susceptible and recovered rate has been found to be inversely proportional, that is the recovered rate has been found to be zero when the susceptible rate has been found to be high and vice-versa. Thus it has been accurately predicted that the recovery rate gradually increased and reached the peak as the days passed.

**Figure.20.**
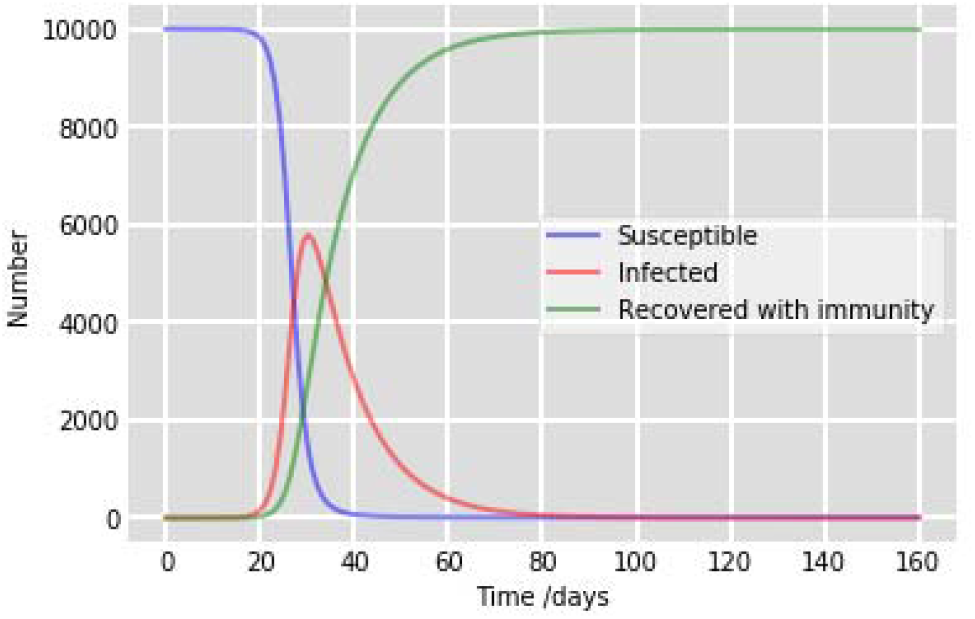
Different cases of COVID-19 in Wyoming.

## V. Conclusion

This proposed study has incorporated the covid-19 pandemic Prediction Model with respect to real pandemic dataset from USA on account of the execution of control measures and the impact of environmental factors to create a prediction system in USA. Based on the SIR model the proposed system is assessed and predicted the covid-19 forecasting in terms of susceptible, recovered and infected in the communities. The prediction of covid-19 with respect to time/days is performed using the modified SIR derived model SIR-D with discrete markov chain. This proposed technique analysed and predicted the covid-19 spread in 19 states of USA.

## Data Availability

N/A

